# Geometric Semi-automatic Analysis of Colles’ Fractures

**DOI:** 10.1101/2020.02.18.20024562

**Authors:** Constantino Carlos Reyes-Aldasoro, Kwun Ho Ngan, Ananda Ananda, Artur d’Avila Garcez, Andy Appelboam, Karen M. Knapp

## Abstract

Fractures of the wrist are common in Emergency Departments, where some patients are treated with a procedure called Manipulation under Anaesthesia. In some cases this procedure is unsuccessful and patients need to visit the hospital again where they undergo surgery to treat the fracture. This work describes a geometric semi-automatic image analysis algorithm to analyse and compare the x-rays of healthy controls and patients with dorsally displaced wrist fractures (Colles’ fractures) who were treated with Manipulation under Anaesthesia. A series of 161 posterior-anterior radiographs from healthy controls and patients with Colles’ fractures were acquired and analysed. The patients’ group was further subdivided according to the outcome of the procedure (successful/unsuccessful) and pre- or post-intervention creating five groups in total (healthy, pre-successful, pre-unsuccessful, post-successful, post-unsuccessful). The semi-automatic analysis consisted of manual location of three landmarks (finger, lunate and radial styloid) and automatic processing to generate 32 geometric and texture measurements, which may be related to conditions such as osteoporosis and swelling of the wrist. Statistical differences were found between patients and controls and pre- and post-intervention, but not between the procedures. The most distinct measurements were those of texture. Although the study includes a relatively low number of cases and measurements, the statistical differences are encouraging.

## Introduction

The dorsally displaced wrist fracture, also known as Colles’ fracture, is the most common fracture of the radius, includes a metaphyseal fracture with a posterior displacement of the distal fragment [1, 2] and can result in some residual impairment in the motion of the hand and wrist [3] and more serious complications such as neuropathies, arthrosis, tendon ruptures and finger stiffness [4]. main procedures for these fractures are Manipulation under Anaesthesia (MUA) and open surgery, also known as Open Reduction and Internal Fixation (ORIF) [5]. MUA, which includes closed reduction and casting, [5], is often the primary option undertaken in Emergency Departments for the displaced fractures in an attempt to correct the deformity and represents a significant proportion of the department workload [6]. Patients are initially treated with a temporary plaster cast after manipulation and a follow-up visit to monitor the rehabilitation progress on a separate day. In general, the fractured position would be improved upon manipulation. There are however cases where the fracture remains unstable or, despite plaster cast immobilisation, slip back into an unacceptable position during rehabilitation. ORIF would then have to be performed with yet another hospital visit causing significant inconvenience to the patient and further inefficiency to hospital resources. ORIF, however, is normally not the preferred option as it requires the booking of operating theatre to operate on the manipulation and fixation of metallic pins, plates or screws. The ORIF procedure is also more complicated than MUA and can lead to serious complications [7].

Despite considerable research [5–11], there is still ambiguity in the procedure to follow with Colles’ fracture [12–14]. There is some evidence that the degree of initial deformity and other factors such as age, dependency, functional status and presence of osteoporosis and x-ray characteristics like axial shortening of bones [15, 16] and angles of volar tilt [17] might predict instability [18–22].

This work describes a geometric semi-automatic image analysis algorithm to analyse and compare the radiographs of healthy controls and patients with Colles’ fractures who have undergone either MUA and were followed to determine if the procedure was successful or unsuccessful. The main objective is to determine if there are geometric differences between the successful and unsuccessful cases. The semi-automatic comparison extracted a series of measurements, e.g. widths of forearm and metacarpal, based on three manually-placed landmarks. In addition, texture measurements were also explored.

## Materials and methods

### Study design and patients

One hundred and sixty-one posterior-anterior radiographs were analysed. Of these, 139 corresponded to wrist fractures and 22 to healthy controls. The wrist fractures were divided by the acquisition time: before (Pre) or after (Post) MUA and the outcome of these: successful or unsuccessful therefore creating four classes pre-successful (n=50), pre-unsuccessful (n=31), post-successful (n=40), post-unsuccessful (n=18). These cases and the clinical outcome were retrospectively identified from electronic attendance logs and electronic records.

The data corresponding to the radiographs that were analysed was anonymised following the ethics procedures at the donating institution and was sourced ethically, with Caldicott Guardian approval, from the Royal Devon and Exeter Hospital. Informed consent was obtained from all individual participants included in the study.

### X-ray acquisition

X-rays were obtained with five different x-ray units: DigitalDiagnost DidiEleva01 (Philips Medical Systems, Netherlands), Mobile tablet work station (Thales, France), DirectView CR 975 and CD 850A (Kodak, USA), Definium 5000 (GE Healthcare, USA) with a variety of exposure factors and saved in DICOM format [23].

Six representative cases of the radiographs are shown in Fig. 1. The radiographs presented considerable variability in the quality, positioning of the arm and presence of collimation lines.

**Fig 1.**
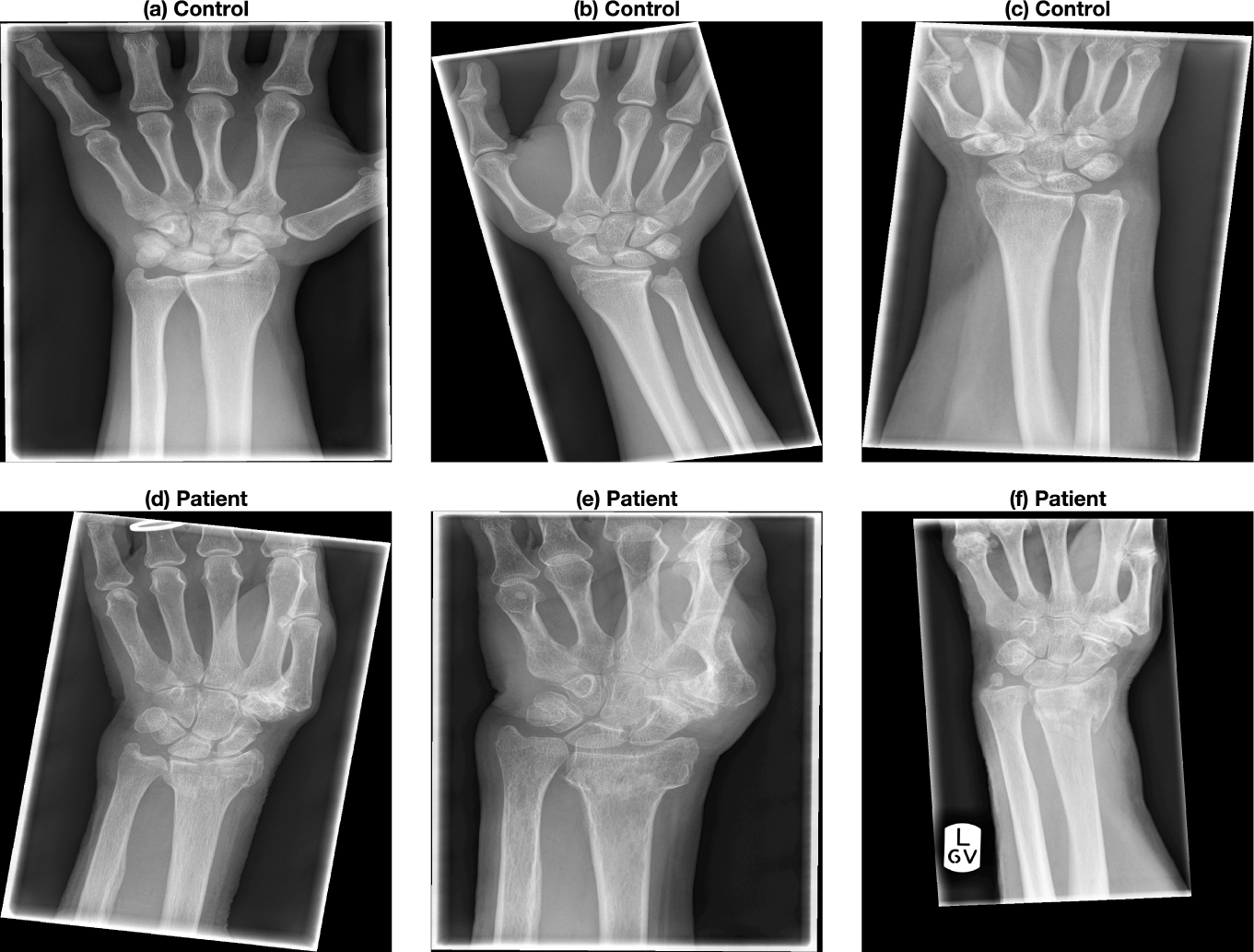
Six representative radiographs that were collected from previous clinical activity at Royal Devon and Exeter NHS Foundation Trust Emergency Department. The images present considerable variability in the quality, positioning of the arm and presence of lines caused by the x-ray collimator. The images were anonymised and metadata such as age, date of acquisition, gender and clinical outcome was available.

### Image analysis

The analysis is considered semi-automatic as three landmarks are manually located and the algorithms obtain all the measurements. All the code was developed in Matlab® (The MathworksTM, Natick, MA, USA) and is available open-source in GitHub (https://github.com/reyesaldasoro/fractures/).

Every image was displayed and three landmarks were manually selected in the following order: (1) base of the lunate, (2) extreme of the radial styloid, (3) centre of the metacarpal of the middle finger (Fig. 3a). These landmarks were subsequently used to obtain a series of measurements described below.

**Fig 2.**
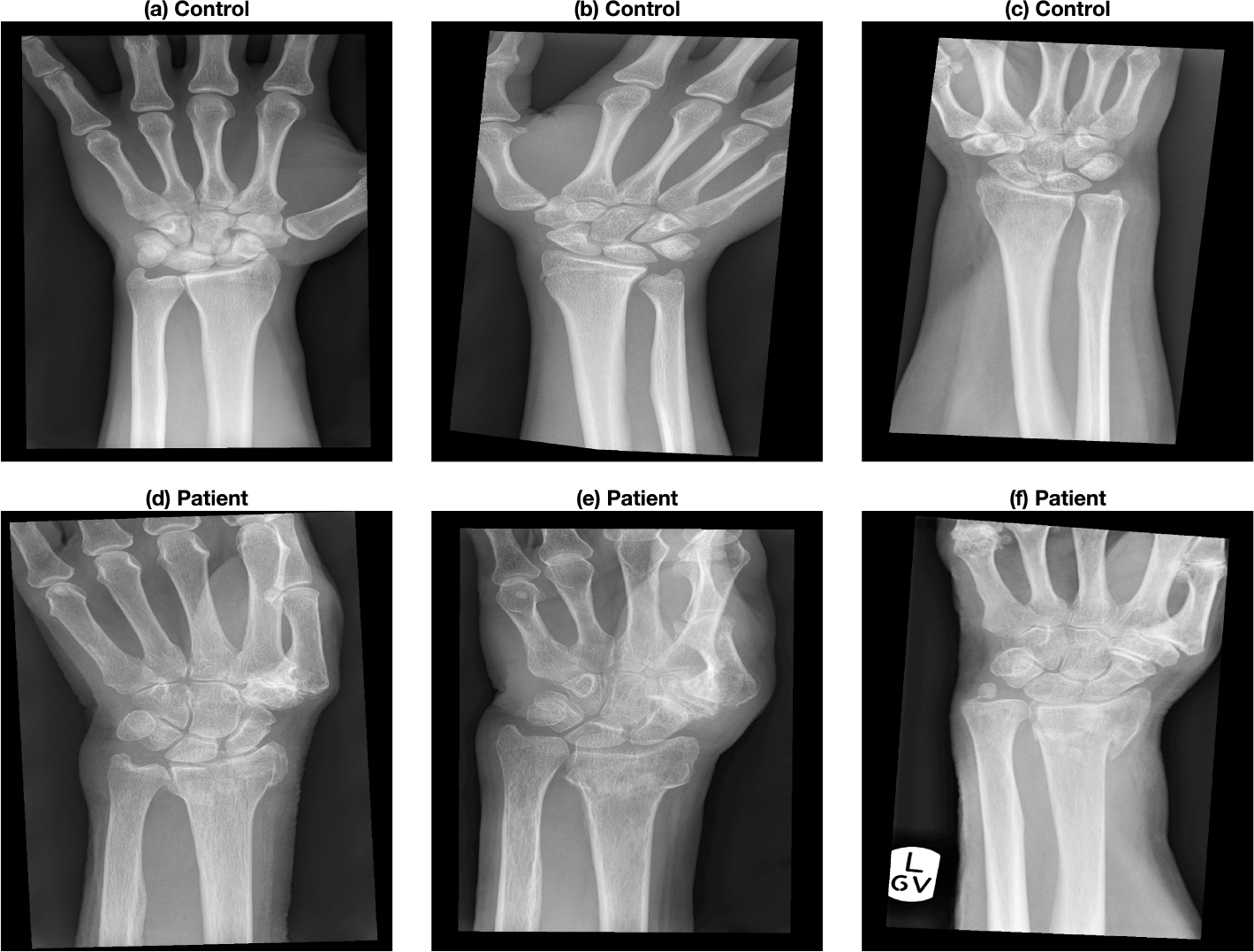
Automatic pre-processing of the radiographs. The six representative cases shown in Fig. 1 were automatically rotated so that the forearm is vertical. In addition, the artefacts due to the collimator were removed. The algorithms were written in Matlab for this proposal and each image is pre-processed in less than one second.

**Fig 3.**
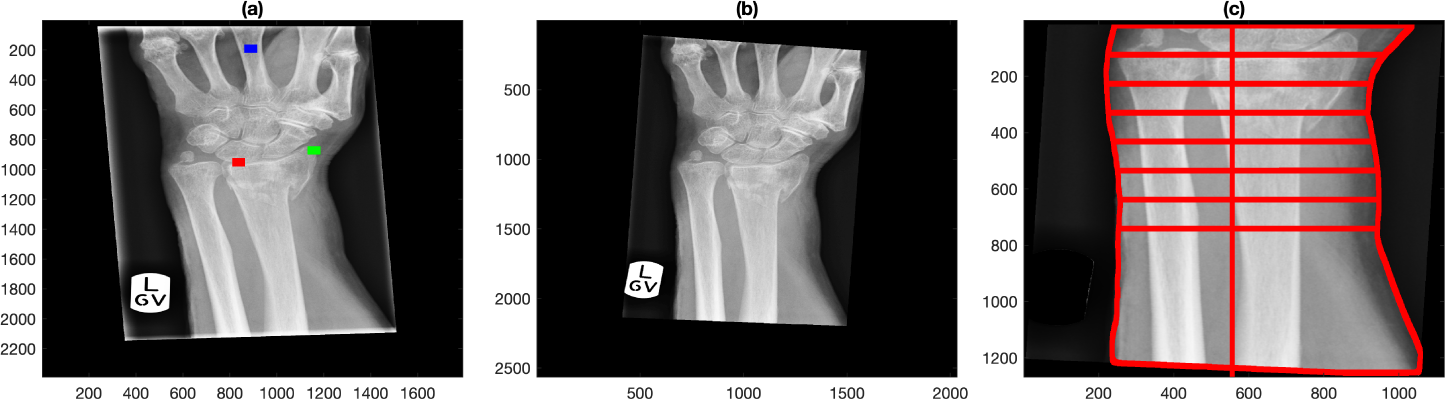
Semi-automatic extraction of measurements of the forearm. (a) Original radiograph that presents rotation of the arm and artefactual lines due to the collimator. Three landmarks have been manually located in the base of the lunate (red), radial styloid (green) and centre of the middle finger (blue). (b) Automatic pre-processing of the image where the forearm was aligned vertically and the lines removed. (c) Using the lunate landmark as a guide, the boundaries of the forearm were automatically delineated and lines traced between the boundaries. The distance between the lines is 1 cm and were being used to derive swelling measurements of the wrist.

The first pre-processing step removed the lines caused by the collimator and then aligned the forearm vertically. For this, the DICOM images and headers were read, converted into Matlab and saved as a *.mat file. The central region of the radiograph was selected by dividing the rows and columns into three thirds and selecting the central part where the bones of the forearm were prominent. Canny edge detection [24] was applied and thus the lines of the bones of the forearm were a good indication of the orientation of the arm. The Hough transform [25] was obtained and the maximum peak was used to determine the rotation required to align the forearm vertically. The lines of the collimator were easily detected as the pixels that were beyond the lines of the collimator were always zero, whilst the darkest regions inside the lines of the collimator, whilst low, were always above zero. Thus, the region(s) outside the lines were detected, dilated and removed from the image. Fig. 2 shows the effect of the pre-processing in the six cases of Fig. 1.

Three groups of measurements were analysed with the expectation that each of these groups would correlate with a clinical condition such as swelling or osteoporosis.

First, as an indication of swelling, the boundaries of the forearm were detected. The landmark of the lunate (Red dot in Fig. 3a) was used to determine the base of the wrist. The region of interest was determined from this point towards the forearm and the region of the hand was removed. The boundaries of the forearm were detected by Canny edge detection and then 8 lines perpendicular to the forearm, each at 1 cm separation were traced. The width of the forearm at each of these lines was recorded (Fig. 3c) with the conjecture that the relationship between the widths could be an indication of swelling of the wrist due to the fracture. A series of ten measurements were generated by calculating ratios, e.g. width at the centre divided by widths at the extremes.

Second, the landmark of the middle finger (blue dot in Fig. 3a) was used to extract a region of interest that contained a segment of the finger (Fig. 4a). The bone in this region was also aligned vertically. Then, the edges of the finger itself and the trabecular and cortical regions were obtained (Fig. 4b) by calculating a vertical projection of the intensities of the image (Fig. 4c). It was conjectured that the thickness of the cortical and trabecular regions of the bone would be an indication of osteoporosis [26–28]. The measurements extracted were the width of the finger and the ratio of trabecular area to total area.

**Fig 4.**
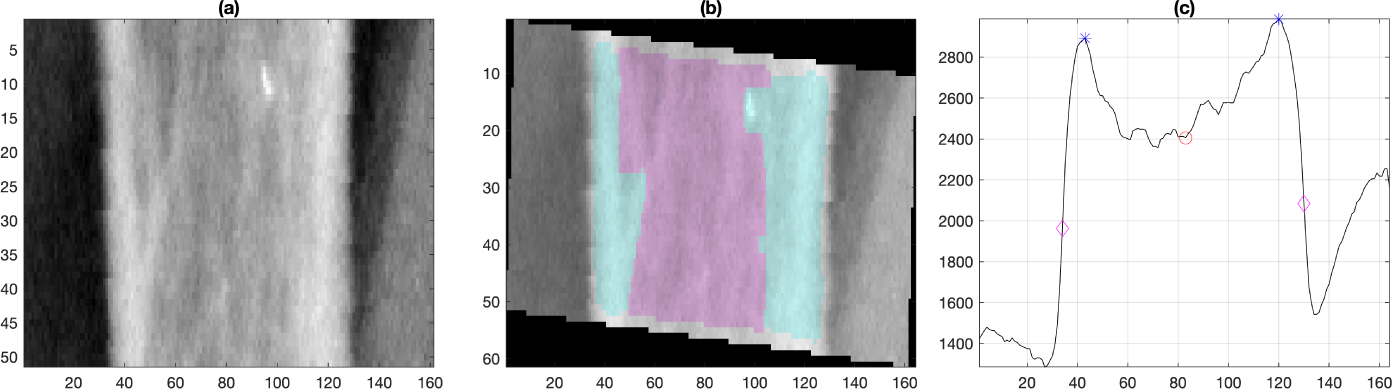
Semi-automatic extraction of measurements of the finger (a) Region of interest (ROI) of the central finger generated from the landmark, blue dot in Fig. 3a. (b) Identification of regions of cortical bone (shaded in cyan) and trabecular bone (shaded in pink) from which the ratio of cortical to total area was calculated. Notice that the finger was rotated to align vertically as the previous rotation aligned the forearm but the fingers are not necessarily vertical. (c) Intensity profile of the ROI with the following key points: edges of the bone (magenta diamond), peak of cortical bone (blue asterisk) and centre of bone (red circle).

Third, preliminary work had identified the potential correlation of texture measurements extracted from x-rays with image analysis with clinical outcome [29]. Therefore, bone texture was analysed in two ways. First, a small region of bone (Fig. 5a,b) was selected from the radius, a small distance away from the landmarks previously detected. This region was analysed with a texture technique called Local Binary Pattern (LBPs) [30], which explores the relations between neighbouring pixels. LBPs concentrate on the relative intensity relations between the pixels in a small neighbourhood and not in their absolute intensity values or the spatial relationship of the whole data. The texture analysis was based on the relationship of the pixels of a 3 *×* 3 neighbourhood. A Texture Unit was calculated by differentiating the grey level of a central pixel with the grey level of its neighbours. The differences were measured if the neighbour is greater or lower than the central pixel and is then recorded as a histogram (Fig. 5c). This analysis provided 10 measurements.

**Fig 5.**
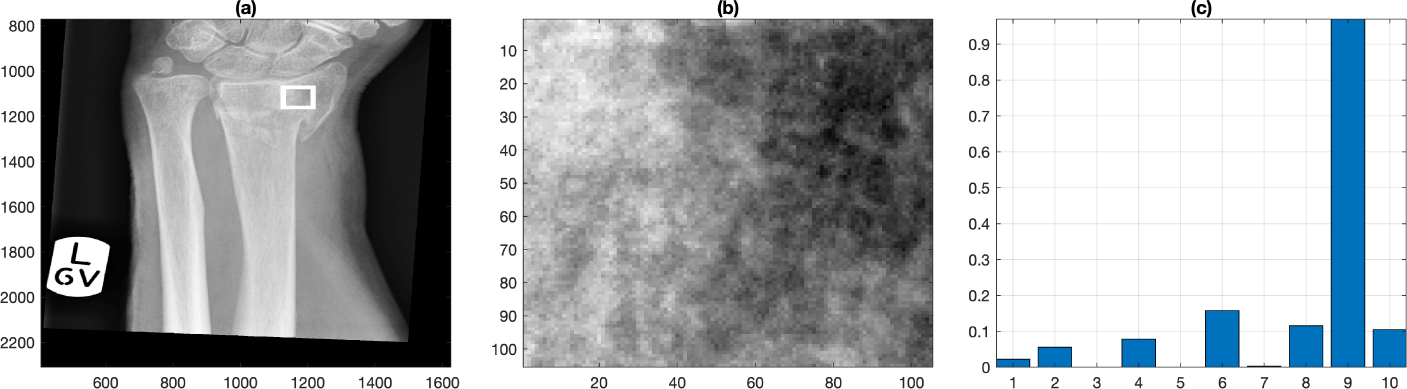
Semi-automatic extraction of texture measurements of a region of interest. (a) To analyse the texture of the radius, a ROI is automatically located by traversing a fixed distance from the radial styloid landmark. (b) Zoom of the region of interest. (c) Texture coefficients generated by Local Binary Pattern analysis.

Another way of analysing the texture of the bones is through intensity profile lines, which capture the variation of the bone intensity over a straight line (Fig. 6a). Initially, a line (green) was automatically traced between the lunate the radial styloid landmarks. Two lines were automatically derived from the first, one at 30 (red) and one at 45 (blue) degrees from the radial styloid up to the edge of the radius. The edge was automatically detected when the intensity dropped drastically into the darker region between the bones. Measurements were extracted both from the intensity profiles (Fig. 6b) and also the profiles after these were adjusted by removing the slope (Fig. 6c) with the idea that measurements like the standard deviation would not be biased by a line that increases its intensity. This analysis provided 10 measurements, e.g. length, slope and standard deviation of the profile.

**Fig 6.**
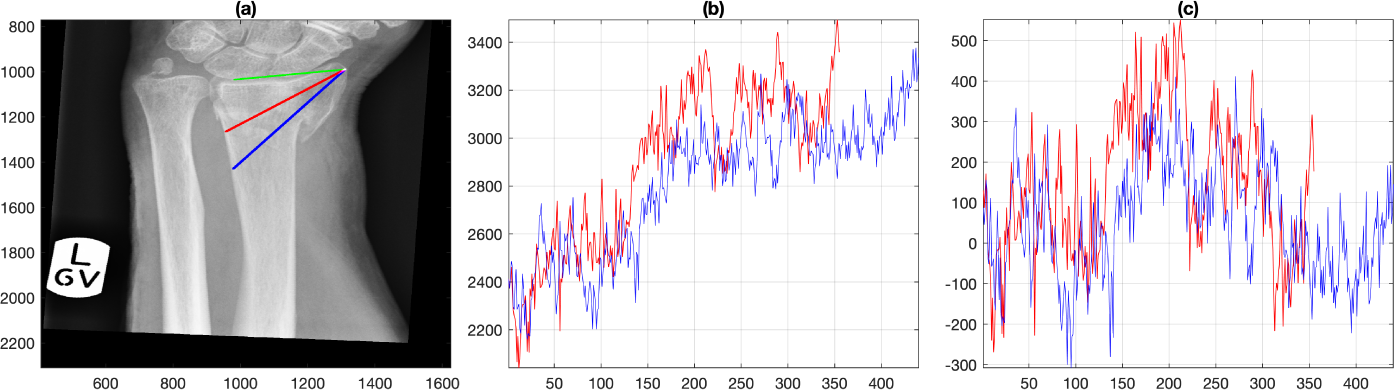
Semi-automatic extraction of texture measurements from intensity profiles. (a) Profile lines from the radial styloid. Initially, a line (green) is automatically traced between the lunate (red in Fig. 3a) the radial styloid (green in Fig. 3a) landmarks. Two lines are automatically derived from the first, one at 30 degrees (red) and one at 45 degrees (blue) from the radial styloid up to the edge of the radius, which is automatically detected. (b) Intensity profiles corresponding to the lines traced in (a). Notice the increasing slope. (c) Intensity profiles adjusted by removing the slope.

All steps except the location of the three landmarks is automatic and takes around 10-20 seconds to process with custom-made Matlab scripts. These scripts are available in the Github repository (https://github.com/reyesaldasoro/fractures/).

## Results

A total of 32 measurements were extracted for each of the radiographs of the five groups previously described and these are presented in Table 1. For each of the measurements, statistical difference between the following cases was tested with paired t-tests: (i) healthy controls against patients. (ii) Pre-intervention (successful and unsuccessful) against post-intervention (successful and unsuccessful). (iii) Successful against unsuccessful. (iv) Pre-intervention successful against pre-intervention unsuccessful. (v) Post-intervention successful against post-intervention unsuccessful. (vi) Pre-intervention successful against post-intervention successful. (vii) Pre-intervention unsuccessful against post-intervention unsuccessful. Three representative measurements are shown in Fig. 7 as boxplots.

**Table 1.** Measurements extracted from the radiographs. Third column corresponds to the landmark used to calculate the measurement. Columns 4-10 show the p-values result of paired t-tests between different groups. Values lower than 0.05 are highlighted in bold. Abbreviations: Ratio of width line 1 / width line 4 (W1 / W4), Local Binary Pattern (LBP), Standard Deviation (Std)

|  | Measurement | Landmark | Control<br>v<br>Patient | Pre-<br>v<br>Post- | Successful<br>v<br>Unsuccessful | Pre-Successful<br>v<br>Pre-Unsuccessful | Post-Successful<br>v<br>Post-Unsuccessful | Pre-Successful<br>v<br>Post-Successful | Pre-Unsuccessful<br>v<br>Post-Unsuccessful |
| --- | --- | --- | --- | --- | --- | --- | --- | --- | --- |
| 1 | W1 / W4 | Lunate | < **0.01** | **0.01** | 0.55 | 0.16 | 0.51 | **0.01** | 0.74 |
| 2 | W2 / W4 | Lunate | < **0.01** | 0.21 | 0.96 | 0.53 | 0.44 | 0.09 | 0.99 |
| 3 | W3 / W4 | Lunate | **0.04** | 0.36 | 0.37 | 0.37 | 0.68 | 0.19 | 0.78 |
| 4 | W5 / W4 | Lunate | 0.10 | 0.30 | 0.38 | 0.19 | 0.33 | 0.94 | 0.09 |
| 5 | W6 / W4 | Lunate | 0.22 | 0.37 | 0.39 | 0.30 | 0.76 | 0.88 | 0.27 |
| 6 | W7 / W4 | Lunate | 0.92 | 0.31 | 0.41 | 0.53 | 0.68 | 0.49 | 0.51 |
| 7 | W8 / W4 | Lunate | 0.45 | 0.25 | 0.65 | 0.50 | 0.77 | 0.54 | 0.33 |
| 8 | Min width /<br>Max width | Lunate | 0.22 | < **0.01** | 0.16 | 0.39 | 0.41 | < **0.01** | 0.11 |
| 9 | W1+W8 /<br>W4+W5 | Lunate | < **0.01** | **0.01** | 0.72 | 0.21 | 0.72 | < **0.01** | 0.53 |
| 10 | W1+W2 /<br>W7+W8 | Lunate | 0.06 | 0.50 | 0.58 | 0.35 | 0.60 | 0.23 | 0.73 |
| 11 | Trabecular<br>Area / Total<br>Area | Finger | < **0.01** | 0.07 | 0.13 | 0.11 | 0.49 | 0.07 | 0.43 |
| 12 | Width Finger | Finger | 0.85 | **0.02** | 0.66 | 0.86 | 0.52 | **0.04** | 0.40 |
| 13 | LBP 1 | L+Rad<br>Sty | **0.02** | **0.01** | 0.28 | 0.96 | 0.07 | **0.01** | 0.54 |
| 14 | LBP 2 | L+Rad<br>Sty | < **0.01** | < **0.01** | 0.41 | 0.83 | 0.09 | < **0.01** | **0.01** |
| 15 | LBP 3 | L+Rad<br>Sty | < **0.01** | < **0.01** | 0.22 | 0.68 | 0.13 | < **0.01** | **0.03** |
| 16 | LBP 4 | L+Rad<br>Sty | < **0.01** | < **0.01** | 0.14 | 0.45 | 0.26 | < **0.01** | < **0.01** |
| 17 | LBP 5 | L+Rad<br>Sty | < **0.01** | < **0.01** | 0.16 | 0.46 | 0.13 | < **0.01** | < **0.01** |
| 18 | LBP 6 | L+Rad<br>Sty | < **0.01** | < **0.01** | 0.08 | 0.24 | 0.30 | < **0.01** | < **0.01** |
| 19 | LBP 7 | L+Rad<br>Sty | < **0.01** | < **0.01** | 0.17 | 0.60 | 0.07 | < **0.01** | < **0.01** |
| 20 | LBP 8 | L+Rad<br>Sty | < **0.01** | < **0.01** | 0.06 | 0.22 | 0.23 | < **0.01** | < **0.01** |
| 21 | LBP 9 | L+Rad<br>Sty | < **0.01** | < **0.01** | 0.15 | 0.57 | 0.11 | < **0.01** | < **0.01** |
| 22 | LBP 10 | L+Rad<br>Sty | < **0.01** | < **0.01** | 0.46 | 0.78 | 0.09 | < **0.01** | **0.01** |
| 23 | Slope profile 1 (full line) | Radial Styloid | < **0.01** | < **0.01** | 0.88 | 0.29 | 0.35 | < **0.01** | 0.09 |
| 24 | Slope profile 2 (full line) | Radial Styloid | 0.04 | < **0.01** | 0.39 | 0.78 | 0.52 | < **0.01** | **0.02** |
| 25 | Slope profile 1 (short segment) | Radial Styloid | < **0.01** | < **0.01** | 0.91 | 0.53 | 0.85 | < **0.01** | < **0.01** |
| 26 | Slope profile 2 (short segment) | Radial Styloid | < **0.01** | 0.06 | 0.82 | 0.74 | 0.85 | 0.11 | 0.29 |
| 27 | Std profile 1 | Radial Styloid | < **0.01** | < **0.01** | 0.92 | 0.39 | 0.59 | < **0.01** | **0.05** |
| 28 | Std profile 2 | Radial Styloid | < **0.01** | < **0.01** | 0.74 | 0.84 | 0.77 | < **0.01** | 0.06 |
| 29 | Std profile 1 adjusted | Radial Styloid | < **0.01** | < **0.01** | 0.50 | 0.80 | 0.47 | < **0.01** | < **0.01** |
| 30 | Std profile 2 adjusted | Radial Styloid | < **0.01** | < **0.01** | 0.10 | 0.42 | 0.22 | < **0.01** | < **0.01** |
| 31 | Distance profile 1 | Radial Styloid | < **0.01** | 0.23 | 0.78 | 0.99 | 0.75 | 0.28 | 0.63 |
| 32 | Distance profile 2 | Radial Styloid | < **0.01** | 0.25 | 1.00 | 0.98 | 0.86 | 0.42 | 0.38 |

**Fig 7.**
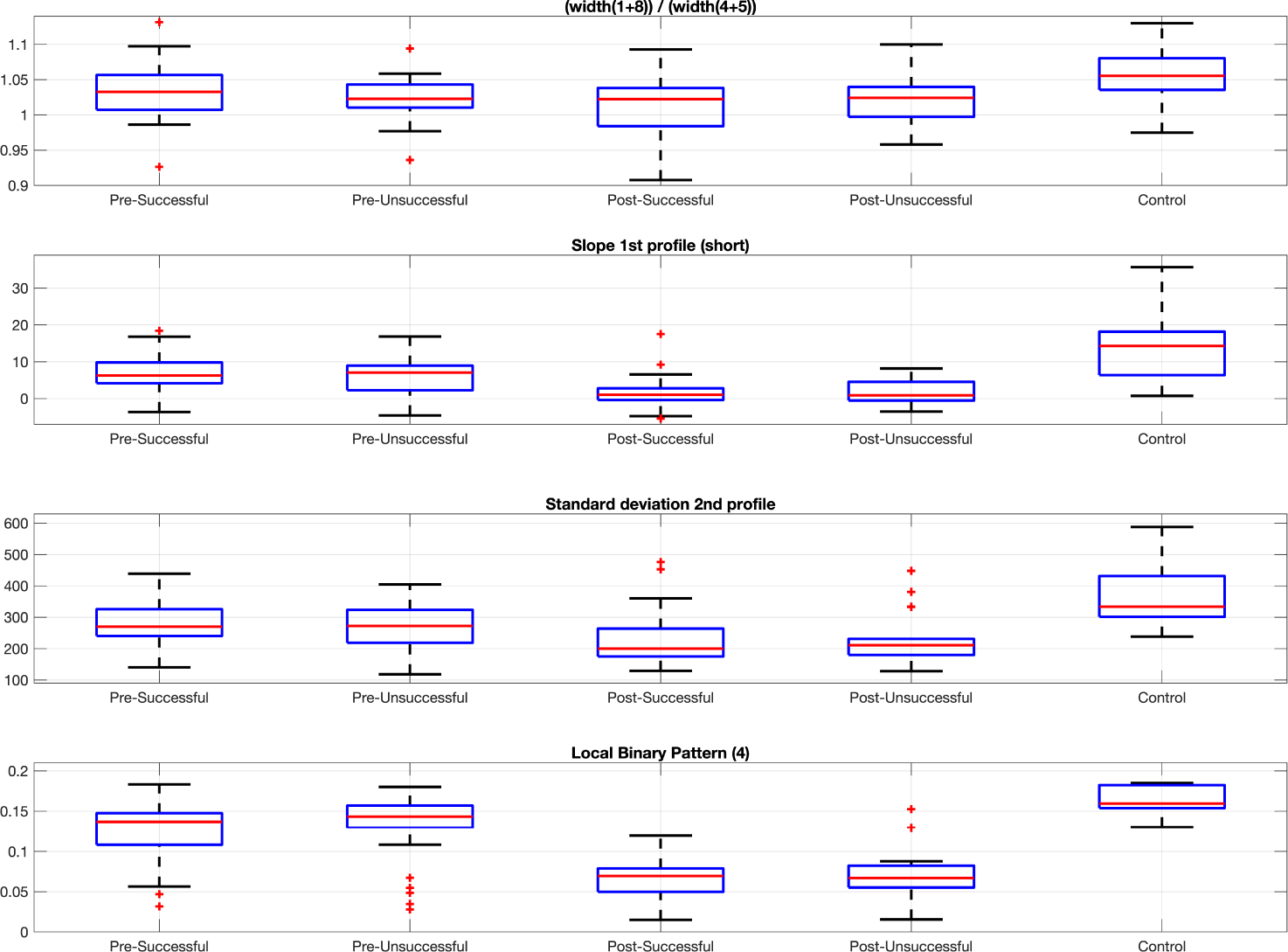
Boxplots corresponding to distributions of four representative measurements. The differences between control and patients and the pre- and post-intervention cases are noticeable but within the pre-intervention and post-intervention groups are very close to each other.

## Discussion

In this work, a series of measurements were extracted from x-ray posterior-anterior images with a semi-automatic methodology. User intervention was minimal and required the selection of three landmarks, which took less than one minute per image. All other steps were automatic and processing of one image took approximately 10-20 seconds. All measurements were validated visually. It should be highlighted that there was a large variation in the quality of the images and this did not affect the measurement extraction. Namely, labels such as those visible in Fig. 3 and the presence of plaster casts did not affect the methodology.

No measurement indicated a statistical difference between the following groups: successful and unsuccessful, pre-intervention successful and pre-intervention unsuccessful, post-intervention successful and post-intervention unsuccessful. However, numerous measurements were statistically different between the groups: healthy controls and patients, pre-intervention and post-intervention for successful, unsuccessful and combined. The differences between healthy controls and patients could be expected due to many factors, among them the age of the patients was higher than the controls. Within the patient groups, the texture features, both those extracted from the profile lines as the LBP features shown statistical difference between controls and patients as well as pre- and post-intervention x-rays showed differentces. Twenty-five of the 32 measurements indicated statistical difference between controls and patients. Similarly, 21 measurements indicated difference between pre- and post-intervention, 21 measurements indicated difference pre-intervention and post-intervention successful and 14 pre-intervention and post-intervention unsuccessful. The LBP measurements were most distinct showing differences for four of the seven groups, followed by the measurements derived from the intensity profiles. These results are encouraging and suggest that the texture features should be further studied, especially analysing the texture in different regions or larger areas as the differences could vary if the location was changed as has been reported in cases of bone mineral density [31] and following for longer periods as the changes in texture are not likely to be changes of osteoporosis given the short time between the pre- and post-imaging.

Whilst none of the results between successful and unsuccessful were significant, some to the texture measurements were close to 0.05. These results are also encouraging, and invite for further experimentation with larger samples and more measurements. New measurements such as radial shortening [32], volar and dorsal displacements [33], ulnar variance, palmar tilt and radial inclination [34] should be explored as these are widely used, but always extracted manually. Similarly, more osteoporosis-related measurements e.g. cortical thickness, internal diameter, cortical area [35] should be explored in regions other than the central finger. Furthermore, the prospect of replacing the semi-automatic nature of the methodology with a fully automatic should be explored. This includes the possible option to incorporate the use of convolution neural networks for automatic detection of the radius [36, 37] and fracture diagnosis [38].

## Data Availability

All the code was developed in Matlab® (The MathworksTM, Natick, MA, USA) and is available open-source in GitHub.

https://github.com/reyesaldasoro/fractures/

## Ethical approval

“All procedures performed in studies involving human participants were in accordance with the ethical standards of the institutional and/or national research committee and with the 1964 Helsinki declaration and its later amendments or comparable ethical standards.”

